# Catecholaminergic modulation of large-scale network dynamics is tied to the reconfiguration of corticostriatal connectivity

**DOI:** 10.1101/2024.07.15.24310279

**Authors:** Justine A. Hill, Cole Korponay, Betty Jo Salmeron, Thomas J. Ross, Amy C. Janes

## Abstract

Large-scale brain network function is critical for healthy cognition, yet links between such network function, neurochemistry, and smaller-scale neurocircuitry are unclear. Here, we evaluated 59 healthy individuals using resting-state fMRI to determine how network-level temporal dynamics were impacted by two well-characterized pharmacotherapies targeting catecholamines: methylphenidate (20mg) and haloperidol (2mg). Network dynamic changes were tested for links with drug-induced alterations in complex corticostriatal connections as this circuit is a primary site of action for both drugs. A randomized, double-blind, placebo-controlled design was used. Methylphenidate enhanced time spent in the default mode network (DMN p<0 .001) and dorsal attention network (DAN p<0.001) and reduced time in the frontoparietal network (p<0.01). Haloperidol increased time in a sensory motor-DMN state (p<0.01). The magnitude of change in network dynamics induced by methylphenidate vs. placebo was correlated with the magnitude of methylphenidate-induced rearrangement of complex corticostriatal connectivity (R=0.32, p=0.014). Haloperidol did not alter complex corticostriatal connectivity. Methylphenidate increased time in networks involved in internal (DMN) and external attention (DAN), aligning with methylphenidate’s established role in attention. Methylphenidate also significantly changed complex corticostriatal connectivity by altering the relative strength between multiple corticostriatal connections, indicating that methylphenidate may shift which corticostriatal connections are prioritized relative to others. Findings further show that these local circuit changes are linked with large scale network function. Collectively, these findings provide a deeper understanding of large-scale network function, set a stage for mechanistic understanding of network engagement, and provide needed information to potentially guide medication use based on network-level effects.

## Introduction

Resting-state brain networks have provided critical insight into the macro-scale functional organization of the brain, which is implicated in healthy cognition and psychopathology(1–3). A key next step is to understand how the function of these distributed brain networks correspond with changes in neurochemistry and more focal functional circuits. While catecholamines, such as dopamine, have been implicated in large-scale network function(3–6), more work using emerging methods is needed to gain a deeper understanding of how catecholaminergic agents impact large-scale network function. Collectively, this line of research promises to enhance the field’s basic neurobiological understanding of brain network function and will shed light on how well-characterized therapeutics impact the brain on a new scale.

A novel approach to assess how catecholamines modulate large-scale network function is network temporal dynamics, which is measured using resting-state functional magnetic resonance imaging (fMRI) data. In contrast to static resting state fMRI analyses which calculates the functional connectivity between brain regions across time, thus leading to one value for the entire resting-state scan, temporal dynamics capture how brain network engagement changes across time. The *resting-state* temporal dynamics of large-scale brain networks have been shown to be central to adaptive functioning, as aberrance in resting-state network temporal dynamics is implicated in numerous forms of psychopathology including ADHD(7,8), nicotine dependence(9–11), cocaine use disorder(12) and schizophrenia(13,14). Dopaminergic transmission has been directly implicated in one *task-based* analysis of network temporal dynamics, which demonstrated that dopamine modulates the dynamics of brain state transitions relevant to working memory(4). However, the evidence linking catecholaminergic transmission to resting-state network dynamics is less clear. One study in children with ADHD indicated that the dopamine and norepinephrine agonist methylphenidate normalized disrupted network temporal dynamics at rest(8). The remaining evidence linking catecholamines and resting-state network temporal dynamics is indirect but widespread, where aberrant catecholaminergic transmission – particularly dopaminergic transmission – is thought to be a principal disruption in many psychopathologies which reliably exhibit altered network dynamics(7–14). Taken together, the direct influence of catecholaminergic transmission on the intrinsic temporal function of the brain’s large-scale networks (e.g., temporal function at rest) remains largely unknown, creating a gap in our understanding of how neuromodulators govern this central component of macro-scale brain function.

Our previous work in healthy young adults from the Human Connectome Project used coactivation pattern analysis (CAP) to define eight transient brain states which include well defined core neurocognitive networks (e.g., the default mode network (DMN), frontoparietal network (FPN), and dorsal attention network (DAN))(15). Here, we apply these state definitions to investigate how acute administration of catecholaminergic agents affect the temporal properties of large-scale brain networks during resting-state fMRI scans in healthy adults. Specifically, in one scan condition we administered methylphenidate (MPH), a dopamine and norepinephrine transporter (DAT/NET) reuptake inhibitor which acts globally to increase extracellular dopamine and norepinephrine and has been shown to enhance cognition due to MPH’s impact on striatal function(16–19). In a second scan condition, we administered haloperidol (HAL), a selective antagonist of D2/D3 receptors located primarily in the striatum. In a third scan condition, we administered placebo. These drugs were administered randomly across participants and allowed us to probe the impact of catecholaminergic agonism (MPH) and D2 antagonism (HAL) on temporal dynamics of large-scale brain networks.

We then move further to investigate ties between catecholaminergic modulation of macro-scale brain network temporal dynamics and more focal circuitry. On the circuit-level, catecholamines, most notably dopamine, are known to act directly on the striatum and alter connectivity between the striatum and the cortex(20). Though most work in fMRI simplifies corticostriatal communication as connectivity between one striatal node and one cortical node, corticostriatal circuitry is much more complex. Striatal nodes project to various cortical regions, while in turn, several cortical regions provide collective and summative input onto striatal nodes(21,22). Preclinical data supports the notion that striatal function is shaped by the convergence of multiple cortical inputs rather than input from any one cortical node alone(21,23). Moreover, the role of striatal dopamine (modulated by both MPH and HAL) may be to alter how *multiple inputs* are *integrated* at striatal cells(20). As such, here we conduct connectivity profile analysis(24) to characterize how MPH and HAL alter multifaceted corticostriatal configuration profiles. We ultimately aim to determine whether the magnitude of catecholamine-driven change in corticostriatal configuration profiles is tied to the magnitude of catecholaminergic modulation of network temporal dynamics. Altogether, our design allows us to establish how dopamine/norepinephrine agonism and D2 antagonism directly impact the inherent temporal function of macro-level brain networks and how this is tied to modulation of specific corticostriatal circuitry.

## Methods and Materials

### Participants

Participants included 59 healthy right-handed individuals between the ages 18-55 (Table 1). Participants were excluded for contraindications with fMRI scanning. Individuals with mental and physical health diagnoses and/or medications that interfere with the bold signal or alter metabolism of catecholaminergic agents were also excluded (See Supplement for details). Participants were recruited from Baltimore, MD, and surrounding areas. The current study was reviewed and approved by the institutional review board of the National Institutes of Health. All participants provided written informed consent.

**Table 1.** Demographics of Sample.

| **Demographics** |  |
| --- | --- |
|  | **Healthy Controls**<br>**(N = 59)** |
| Age (years) | 39.32 (*SD* = 11.25) |
| Sex (male/female) | 17/42 |
| Race (*n*) |  |
| Black/African American | 11 |
| White/Caucasian | 37 |
| Asian | 5 |
| More Than One Race | 6 |
| Ethnicity (*n*) |  |
| Hispanic | 9 |
| Non-Hispanic | 49 |
| Unknown or not reported | 1 |
| Education (*n*) |  |
| Less than High School | 2 |
| High School Complete/GED | 8 |
| Some / Partial Post-High School | 22 |
| College Graduate/Bachelor's Degree | 18 |
| Master's Degree | 5 |
| Professional Degree (MD, JD, Ph.D.) | 4 |

Demographic characterization of sample.

### Study Design

All participants underwent resting-state fMRI scanning sessions with 3 drug conditions administered on separate days, in a double-blind placebo-controlled manner: placebo/placebo, HAL/placebo, placebo/MPH. Doses were 2 mg oral HAL and 20 mg oral MPH. Each scanning visit was identical and took place at drug peak: 4-hours post HAL/placebo and 1-hour post MPH/placebo; timed in accordance with absorption rates to ensure high and stable plasma levels of medication during the scan. Prior to any medication administration, participants completed a nursing assessment, and following the scan session, participants met again with nursing staff to assess side effects.

### Neuroimaging Data

At each scan, data were collected using a Siemens Trio 3T scanner with a 12 channel RF coil. For high-resolution anatomical scan, multi-planar rapidly acquired gradient echo-structural images were obtained with the following parameters (TR=1.9s, TE= 3.51ms, slices=208, matrix=192×256, flip angle 9°, resolution 1.0×1.0×1.0 mm). For the 8-minute resting state scan, gradient-echo, echo planar images were collected using oblique axial scans 30° from AC-PC with AP phase encoding and the following parameters (TR=2s, TE=27ms, flip=78°, vox resolution=3.4375×3.4375×4 mm). Total frames were 240 and the first 5 were discarded.

Images were processed using FMRIB software library (FSL 6.0.0). Images underwent brain extraction, registration, spatial smoothing (6mm), high pass temporal filtering (100s), and motion correction via MCFLIRT. Data were further cleaned to reduce motion artifacts using Independent Component Analysis via FIX(25,26) through MELODIC. Data were then standardized to MNI152 brain image using FLIRT.

### Pharmacological Impact on Network Temporal Dynamics

Previously, Janes et. al(15) performed Co-Activation Pattern Analysis (CAPs) on resting state data from 462 individuals in the Human Connectome Project(27) using 129 ROIs (cortical and striatal ROIs based on functional parcellation(28,29); amygdala ROIs based on anatomical parcellation(30)) and determined eight co-activation patterns (brain states) using k-means clustering. These eight brain states align with previously established resting state networks such as: the DMN, FPN, DAN, SN, and sensorimotor network (SMO) (Supplementary Figure 1). To investigate how these eight brain states behave dynamically at rest under different catecholaminergic drugs, we extracted ROI time courses from the 129 ROIs and conducted CAPs using the Capcalc package (https://github.com/bbfrederick/capcalc). The eight states were used to compute state-specific dynamic measures including: 1) *total time* spent in each state, 2) number of *transitions* (i.e. entries) into each state, 3) average *persistence* within the state once a transition into the state had occurred.

To determine an effect of drug on temporal dynamics of brain state activity at rest, we ran a repeated measures ANOVA that tested the drug × brain state interaction on total time.

Following a significant drug × brain state interaction, we ran eight subsequent repeated measures ANOVAs, testing effect of drug on total time in each state. In states where there was a significant drug effect on total time spent in brain state after correcting for multiple comparisons (Bonferroni corrected: .05/8=.00625), two post-hoc paired t-tests compared MPH and HAL to placebo (Bonferroni corrected: .05/2=.025). All ANOVAs and post-hoc t-tests were conducted in R using the following packages: lme4 (1.1-32), lmertest (3.1-3), effectsize (0.8.3), and multcomp (1.4-23). All analyses controlled for age and sex.

In brain states which exhibit significant change in total time in state under drug, we investigated whether changes to total time in state were driven by changes in 1) number of transitions to the state or 2) persistence within state. We ran separate post-hoc paired t-tests comparing 1) transitions and 2) persistence between drug of interest to placebo. Post-hoc analyses were Bonferroni corrected to account for all states tested. See supplementary methods for detailed description of transition analysis.

### Ties between Network Temporal Dynamics and Static Network Function

Previous work has theorized that spending more time in the DMN and DAN at opposing times drives enhanced static DMN-DAN anti-correlation(31) – a known marker of healthy cognition(32,33). Separate literature consistently shows that MPH enhances static DMN-DAN anti-correlation(34,35). We aimed to confirm that MPH enhances static DMN-DAN anti-correlation in our sample and tested the conjecture that static DMN-DAN anti-correlation is driven by DMN and DAN temporal dynamics. To do so, we obtained DMN-DAN anti-correlation values by regressing the whole brain 4D data on the spatial patterns of DAN and DMN states using MATLAB 2022b. Correlation values between each network’s time course were computed and Z-transformed using Fisher’s R-to-Z. We then tested effect of drug on DMN-DAN anti-correlation with a repeated measures ANOVA and post-hoc t-tests (Bonferroni corrected). Next, we tested the relationship between static DMN-DAN anti-correlation and combined time spent in DMN and DAN, hypothesizing that increased time in DMN and DAN would be associated with greater magnitude of DMN-DAN anti-correlation (based on previous proposal(31)). We fit a linear mixed model in R to estimate DMN-DAN anti-correlation score from time in DMN and DAN state combined (summed), when controlling for age, sex, drug, and drug × time in state interaction.

### Pharmacological Impact on Corticostriatal Configuration

To detect complex changes in corticostriatal connectivity, we conducted connectivity profile analysis(24). Striatal function is shaped by the convergence of multiple cortical inputs more so than by any one cortical node alone(21,23). Here, connectivity profile analysis allowed us to model the multifaceted input received by striatal nodes from diverse areas of cortex, and study drug-related changes in the properties of these “corticostriatal configuration profiles” (CSCPs)(24).

To derive CSCPs, time courses were first extracted from each voxel of a striatal mask(24) and from 53 cortical ROIs (used in CAPs states(15)). Correlation values between time courses at each striatal voxel and each cortical ROI were calculated, then r values were Z-transformed using Fisher’s R-to-Z. Next, CSCP metrics were calculated in MATLAB (2022b) (For details, see Korponay et al (2022), and Supplement). Briefly, CSCP metrics include 1) *aggregate divergence* (AD): absolute sum of the change in connectivity for each ROI at each striatal voxel (drug – placebo), acting as a measure of absolute change in magnitude of connectivity under drug; 2) *rank order rearrangement* (ROR): the absolute sum of change in order of strength of connectivity for each ROI at each striatal voxel (drug – placebo), acting as a measure of absolute change in relative connectivity under drug; 3) e*ntropy shift* (ES): change in distribution of the strength of corticostriatal connectivity values under drug compared to placebo. Here, CSCPs were conducted entirely within-subject. Because CSCP metrics inherently compare drug to placebo, we also conducted within-session placebo CSCP metrics for statistical analysis (PBO_firsthalf_-PBO_secondhalf_). This score compared the first and second half of placebo data. In total, scores were derived across three conditions: MPH-PBO, HAL-PBO, PBO_firsthalf_-PBO_secondhalf_.

To determine statistical significance, we calculated subject-wise average scores of each CSCP metric for each condition. We then ran three repeated measures ANOVAs – one for each metric (AD, ROR, ES) – comparing the drug conditions (Bonferroni corrected). If significant, post-hoc t-tests compared MPH-PBO, HAL-PBO and PBO_firsthalf_-PBO_secondhalf_ (Bonferroni corrected). Where post-hoc t-test revealed a significant drug effect, we then aimed to identify specific areas of the striatum which were significantly altered by drug. We computed subject-level striatal maps of significant CSCP metrics. We then performed a voxel-wise paired t-test between significant drug-placebo and within-session placebo condition using SPM12.

### Ties between Modulation of Network Temporal Dynamics and Corticostriatal Reconfiguration

We tested whether, under the same drug, significant change in network temporal dynamics was related to significant alterations in CSCP metrics. To do so, first we calculated one subject-wise score of absolute change in time spent in states under drug. Absolute change in time spent in states was derived by summing the absolute value of change in time in each of the eight states (time_stateN_ under drug – time_stateN_ under placebo). We then tested the relationship between subject-wise score of significant CSCP metric and absolute change in time spent in states. Where there was a significant relationship, we aimed to confirm that this relationship, involving change in time spent in *all* states, was specifically driven by change in time spent in states whose temporal dynamics were determined to be significantly modulated by drug. The goal of this confirmation was to parse out which networks are involved in the relationship between temporal dynamic network properties and functional corticostriatal circuitry. This was assessed by comparing correlations(36) where x remained CSCP score while y varied as absolute change in: total time spent in all eight states, total time spent in states significantly altered by drug, and total time spent in states not significantly altered by drug. See exact equations in supplementary methods.

We conducted further exploratory post-hoc analysis to localize which striatal nodes and individual networks are involved in significant relationships. In contrast to a CSCP score defined as the mean CSCP score at *all* striatal voxels per subject, node-specific CSCP scores were derived from the mean of voxels in each significant striatal node identified in the voxel-wise t-test. Correlations were calculated between node-specific CSCP scores and change in time spent in each state significantly modulated by drug.

## Results

### Network Temporal Dynamics

We first tested for effect of drug on total time spent in brain states. There was a significant effect of brain state (F(16,928)=19.53, p<0.001) and a drug × brain state interaction (F(16,928)=5.78, p<0.001) on total time spent in brain states. A significant effect of drug was noted for the DMN (F(2,116)=15.29, p_corr_<0.001, _η_^2^=0.21), DAN (F(2,116)=12.42, p_corr_<0.001, _η_ =0.18), FPN (F(2,116)=5.95, p_corr_<0.05, _η_ =0.09) and sensorimotor-occipital DMN (SM-DMN) state (F(2,116)=9.97, p_corr_<0.001, _η_^2^=0.15).

Post-hoc analysis revealed that relative to placebo, MPH increased time spent in the DMN (p_corr_<0.001, d=0.74) and DAN states (p_corr_<0.001, d=0.665). Increased time was explained by MPH increasing the number of transitions into the DMN (p_corr_<0.001, d=0.72) and DAN states (p_corr_<0.001, d=0.78). In contrast, MPH had no impact on persistence in either state (p_corr_>0.05). Given that the DMN and DAN play substantial roles in internal(37–40) and external attention(37,41), respectively, this finding builds on previous literature of MPH’s known role in attentional enhancement and provides novel insight whereby MPH inherently increases transitions into attention-related networks. MPH also reduced the time spent in the FPN state relative to placebo (p_corr_<0.01, d=-0.445) and reduced transitions into this state at a significance level that did not withstand Bonferroni correction (p_corr_=0.17) without influencing persistence (p_corr_>0.05) (Figure 1A-C).

**Figure 1.**
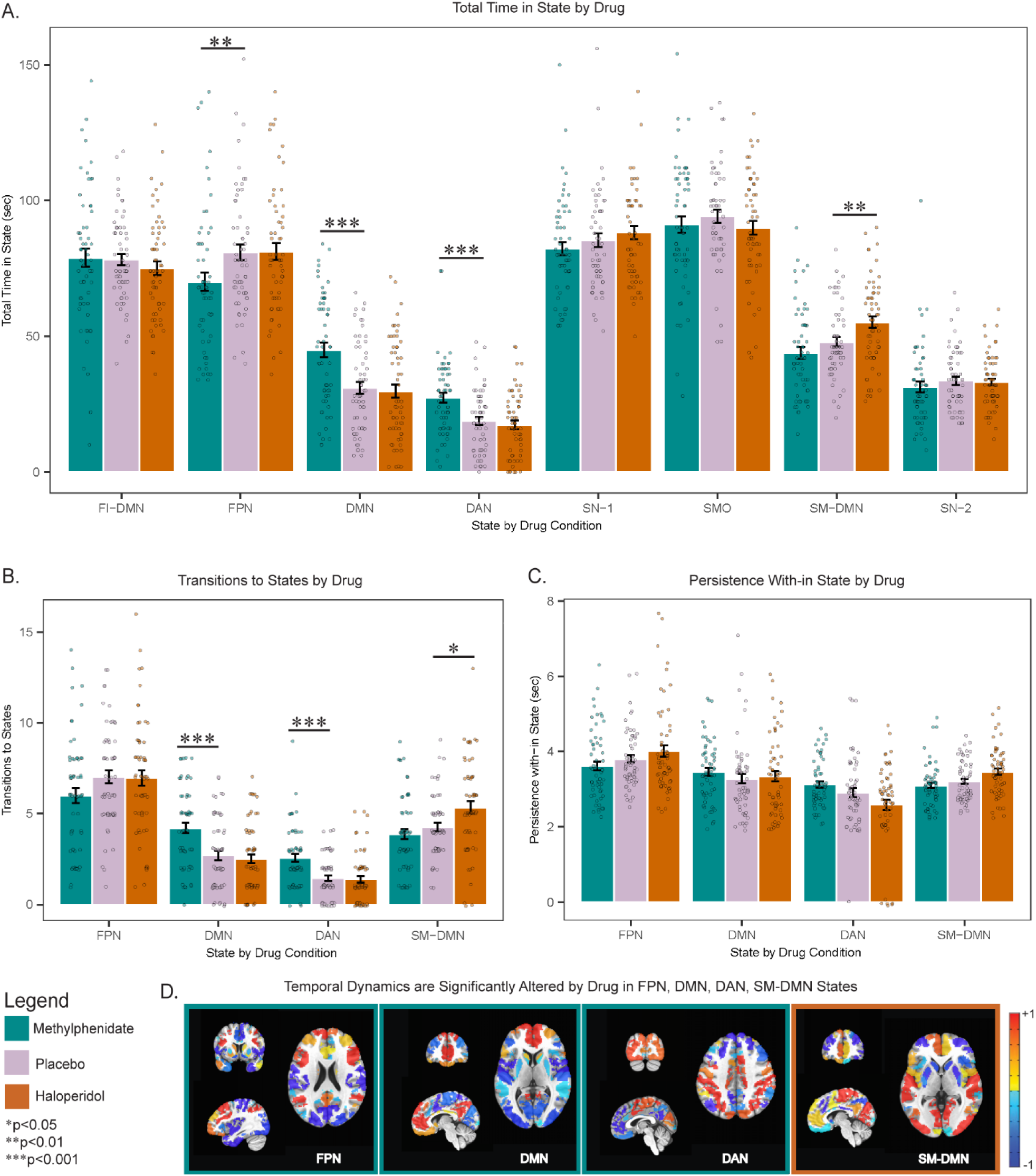
Catecholaminergic Agents Significantly Alter Large-Scale Brain Network Temporal Dynamics. A. MPH increased time spent in DMN and DAN states and decreased time spent in FPN state. HAL increased time spent in SM-DMN state. B. MPH increased transitions to DMN and DAN states while HAL increased transitions to SM-DMN state. C. Neither MPH nor HAL altered time spent persisting within DMN, DAN, FPN states or SM-DMN state. D. A visualization of FPN, DMN, DAN and SM-DMN states of which dynamics are significantly altered by MPH (outlined in green) or HAL (outlined in orange). -1 is most negative relative activation; +1 is most positive relative activation. MPH: methylphenidate; HAL: haloperidol; FI-DMN: fronto-insular default mode network state; FPN: frontoparietal control network state; DMN: default mode network state; DAN: dorsal attention network state; SN-1: salience network state 1; SMO: sensory motor occipital state; SM-DMN: sensory motor default mode network state; SN-2 salience network state 2. *p_corr_<0.05, **p_corr_<0.01, ***p_corr_<0.001

Relative to placebo, HAL increased the time spent in the SM-DMN, which represents the co-activation of sensory motor regions and the DMN (p_corr_<0.01, d=0.49). HAL increased the number of transitions into this state (p_corr_=0.013, d=0.48) and increased the persistence within the SM-DMN at a level that did not withstand Bonferroni correction (p_corr_=0.054) (Figure 1A-C). The effect of HAL on sensory motor-related state dynamics fits with previous findings demonstrating that HAL impacts sensory motor systems(42) and demonstrates that MPH and HAL impact the inherent temporal function of different large-scale networks.

### DAN / DMN Anti-correlation

There was a significant effect of drug on the DMN-DAN anti-correlation (F(2,116)=11.6, p<0.001, ^2^**=**0.17). Post-hoc comparisons confirmed that MPH had a significantly greater magnitude of static DMN-DAN anti-correlation versus placebo (Figure 2A, p_corr_=0.0014, d=0.57) which aligns with previous literature(34,35), whereas HAL did not significantly differ from placebo (p_corr_>0.05). We then tested the hypothesis that greater magnitude of static DMN-DAN anti-correlation could be driven by increased time spent in both the DMN and DAN. Time in DMN and DAN combined negatively estimated DMN-DAN correlation independently of drug conditions (Figure 2B, p<0.001, conditional R^2^=0.72, marginal R^2^=0.59). This lends significant evidence towards the conjecture(31) that enhanced anti-correlation of the DMN and DAN is driven by spending more time both states, at opposing times.

**Figure 2.**
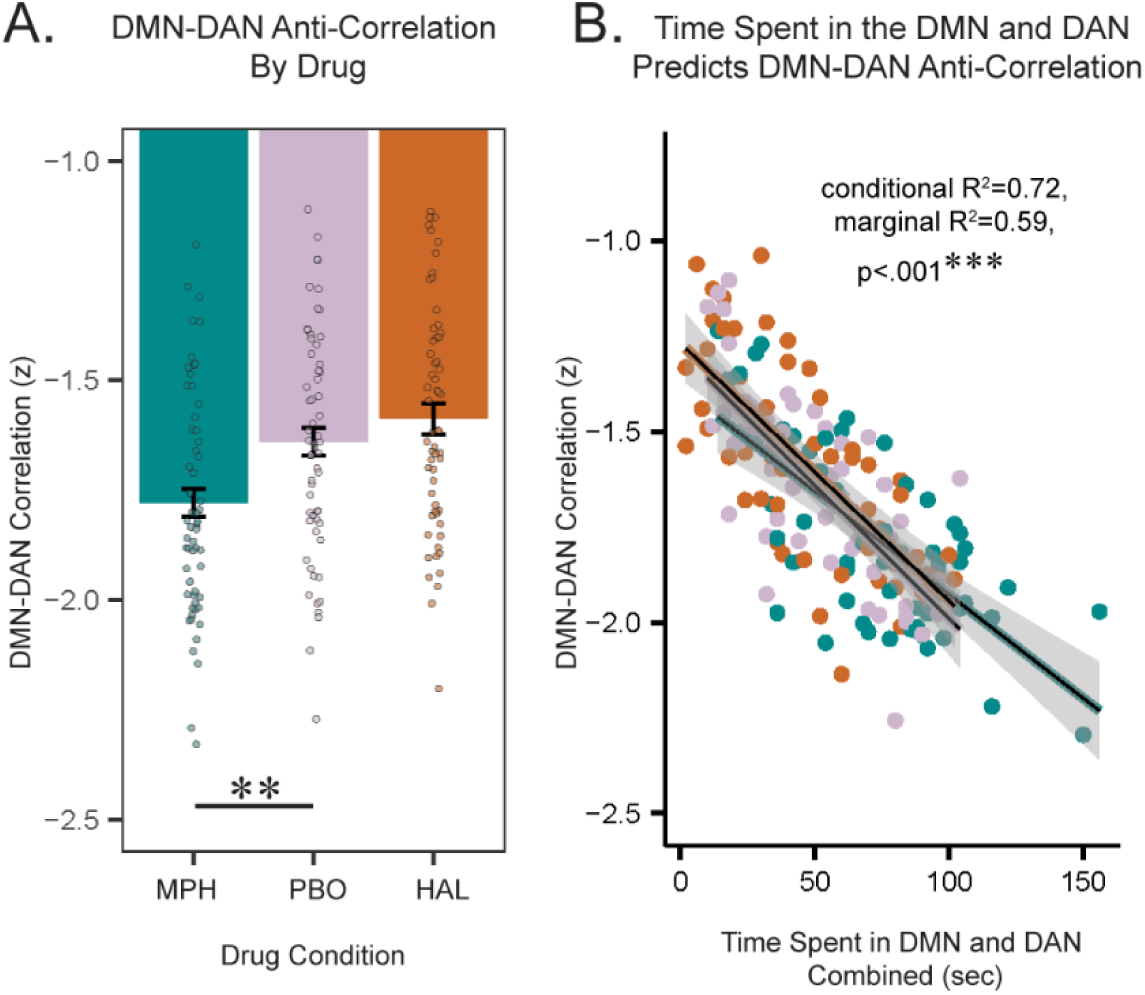
DMN-DAN Static Anti-Correlation is Tied to DMN and DAN Temporal Function. A. MPH significantly strengthens DMN-DAN anti-correlation. B. The combination of time spent in DMN and DAN negatively predicts DMN-DAN correlation values independently of drug. MPH: methylphenidate; HAL: haloperidol; DMN: default mode network; DAN: dorsal attention network. **p_corr_<0.01

### Corticostriatal Configurations

We measured catecholaminergic manipulation of corticostriatal circuitry by measuring effect of drug on CSCP metrics: AD, ROR, and ES. There was a significant effect of drug on ROR, the measure of absolute change in *relative* corticostriatal connectivity, following repeated measures ANOVA (F(2,116)=15.36, p_corr_<0.001, _η_**^2^**=0.21). There was no effect of drug on AD or ES (p_corr_>0.05), which measure the absolute magnitude of change in corticostriatal connectivity values and change in distribution of the strength of corticostriatal connectivity values, respectively. Post-hoc comparisons revealed that the MPH-PBO condition exhibited significantly higher ROR compared to within-session placebo (p_corr_<0.001, d=0.86) and HAL-PBO (p_corr_<0.001, d=0.73; Figure 3A). There was no difference between HAL-PBO and within-session placebo (p_corr_>0.05). This result shows that corticostriatal connectivities which were relatively weaker under placebo became relatively stronger under methylphenidate and vice versa, at a statistically significant level when measuring total amount of change in relative strength. Thus, we provide evidence that elevated extracellular dopamine and norepinephrine alter which specific corticostriatal connections are the strongest or weakest, where this “*reshuffling*” of the relative connectivity strength of cortical regions with the striatum potentially reflects a shift in the priority of corticostriatal communications.

**Figure 3.**
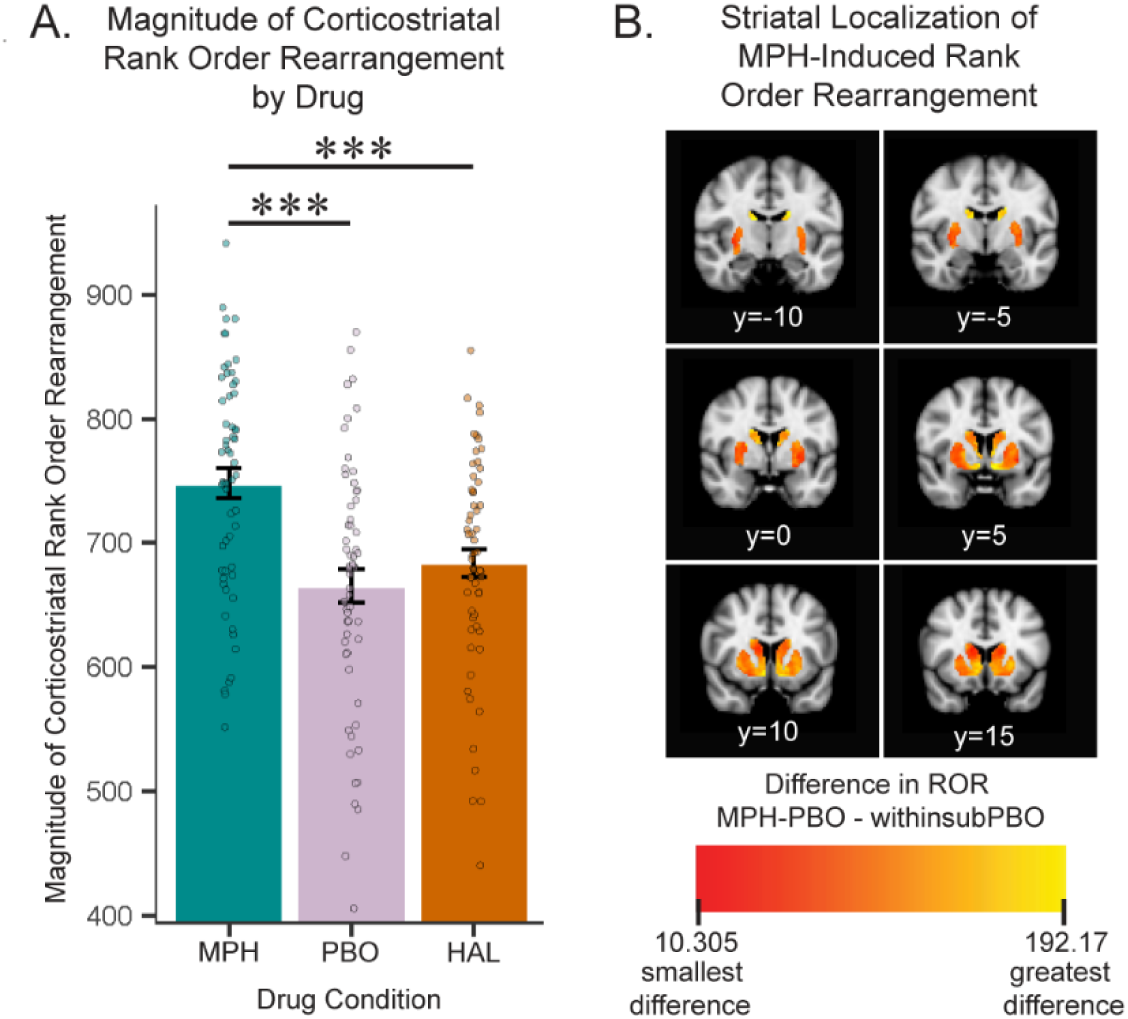
Methylphenidate Significantly Heightens the Magnitude of Rank Order Rearrangement in Corticostriatal Connectivity Profile. A. Under MPH, there is significantly higher magnitude of ROR – measure of absolute change in relative corticostriatal connectivity - compared to placebo (within-session placebo rearrangement) and HAL (HAL-PBO rearrangement). B. Heatmap comparing magnitude of ROR at striatal voxels in MPH-PBO condition compared to within-session placebo condition. Across PBO and MPH conditions, relative connectivity strength between multiple cortical regions and the more **red** striatal voxels is more stable. In contrast, more yellow striatal voxels exhibiter greater change in relative connectivity strength with cortical regions under MPH compared to PBO. ROR: rank order rearrangement; MPH: methylphenidate; HAL: haloperidol; PBO: placebo. ***p_corr_<0.001

To localize striatal nodes where MPH significantly induced higher magnitude of ROR compared to placebo, we conducted a voxel-wise paired t-test of striatal ROR value maps comparing MPH-PBO to within-session placebo. This revealed five clusters where MPH significantly induced higher magnitude of ROR compared to placebo: right dorsal caudate; left dorsal caudate; right nucleus accumbens and ventral caudate; left nucleus accumbens; left ventral caudate (Supplementary Table 1). See supplementary results for details on how cortical regions changed in rank order at significant striatal clusters.

### Network Temporal Dynamics **×** Corticostriatal Configurations

To determine links between smaller-scale circuitry and the time-varying engagement of large-scale networks, we tested associations between significant corticostriatal reconfiguration and change in network temporal dynamics under the same drug. MPH both significantly increased the absolute change in relative corticostriatal connectivity (ROR) and altered network temporal dynamics (Figure 1 and 3). We thus tested the association between magnitude of ROR and change in time spent in network-aligned brain states under MPH. There was a significant positive relationship between magnitude of ROR (averaged across the entire striatum) and absolute change in time spent in all brain states under MPH (R=0.29, p=0.025).

We examined whether this relationship was driven by change in time spent in the FPN, DMN and DAN states, as we determined that MPH significantly altered the time spent in these three states (Figure 1A). There was a significant relationship between magnitude of ROR and absolute change in time spent in FPN, DMN and DAN states under MPH (R=0.32, p=0.014) while there was no significant relationship between magnitude of ROR and absolute change in time spent in all states excluding FPN, DMN and DAN (R=0.12, p>.05) (Figure 4). The relationship between magnitude of ROR and absolute change in time spent in all states was significantly stronger than the relationship involving change in time spent in states *excluding* FPN, DMN, DAN states (z=1.87, p=0.031) and not significantly different from the relationship involving only absolute change in time spent in FPN, DMN and DAN states (z=0.37, p>0.05) (Figure 4). We thus concluded that MPH-induced change in time spent in FPN, DMN and DAN combined is principally responsible for the association between MPH-induced change in time spent in all states and magnitude of corticostriatal ROR.

**Figure 4.**
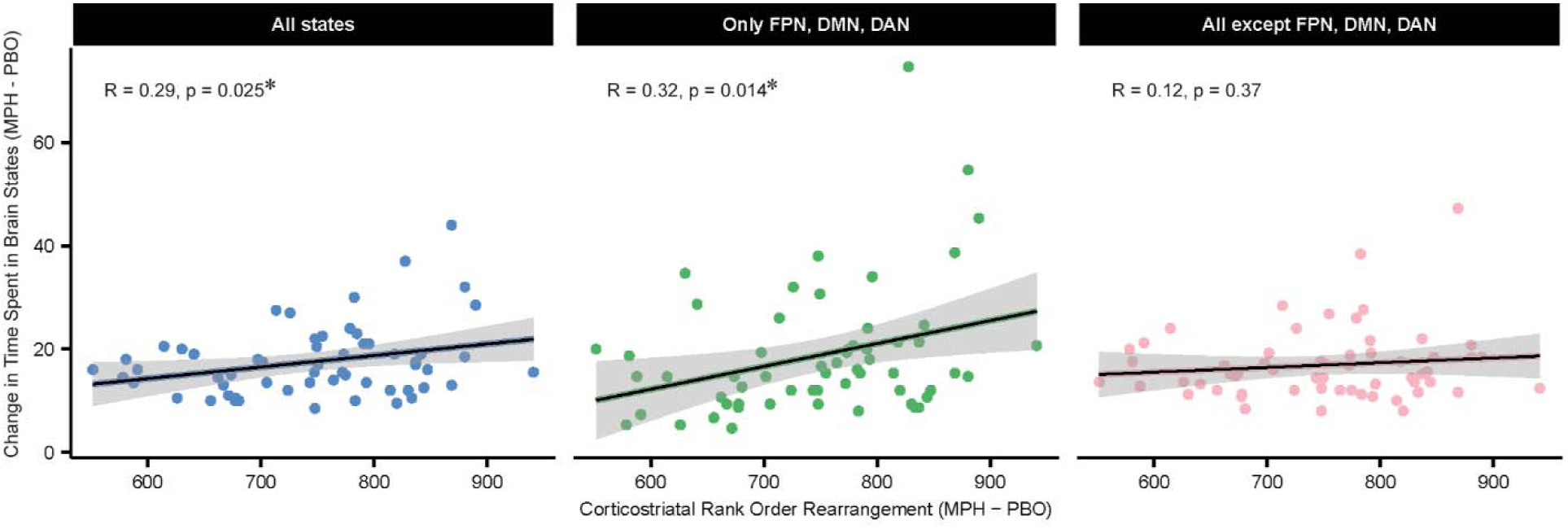
Change in Network Temporal Dynamics is Related to Modulation of Functional Corticostriatal Circuitry under Methylphenidate. Change in network temporal dynamics is associated with altered corticostriatal rank order rearrangement under MPH. Change in network temporal dynamics is operationalized as the sum, for each state tested, of the absolute value of the difference in total time spent in the state (MPH-PBO). Three panels vary by y-axis where left panel y = change in time spent in all states, middle panel y = change in time spent in only FPN, DMN and DAN states, right panel y = change in time spent in all states except FPN, DMN, and DAN states. All Y-axis measures were divided by the number of states summed. MPH: methylphenidate, PBO: placebo; DMN: default mode network; DAN: dorsal attention network; FPN: frontoparietal network.

In an exploratory analysis, we aimed to localize specific striatal nodes where magnitude of ROR may be associated with change in time spent in the FPN, DMN or DAN. We averaged ROR values for each subject at each of the five striatal nodes previously identified (Supplementary Table 1). We then tested associations between node-wise ROR and change in time spent in FPN, DMN and DAN independently – ultimately yielding 15 correlation tests (5 nodes × 3 brain states). Change in time spent in the DMN alone was significantly positively related to ROR at the left dorsal striatum (Figure 5B, R=0.4, p_uncorr_=0.0015; p_corr_=0.0225). The positive relationship with the right dorsal striatum (Figure 5D, R=0.35, p_uncorr_=0.006, p_corr_>0.05) did not survive multiple comparisons correction. However, these relationships did not significantly differ from each other (z=-0.87, p>0.05). No other relationships were significant following multiple comparisons correction. These exploratory findings demonstrate that the magnitude at which MPH reshuffles the relative connectivity strength between different cortical regions and the dorsal caudate is associated with the amount that MPH enhances time spent in the DMN at rest.

**Figure 5.**
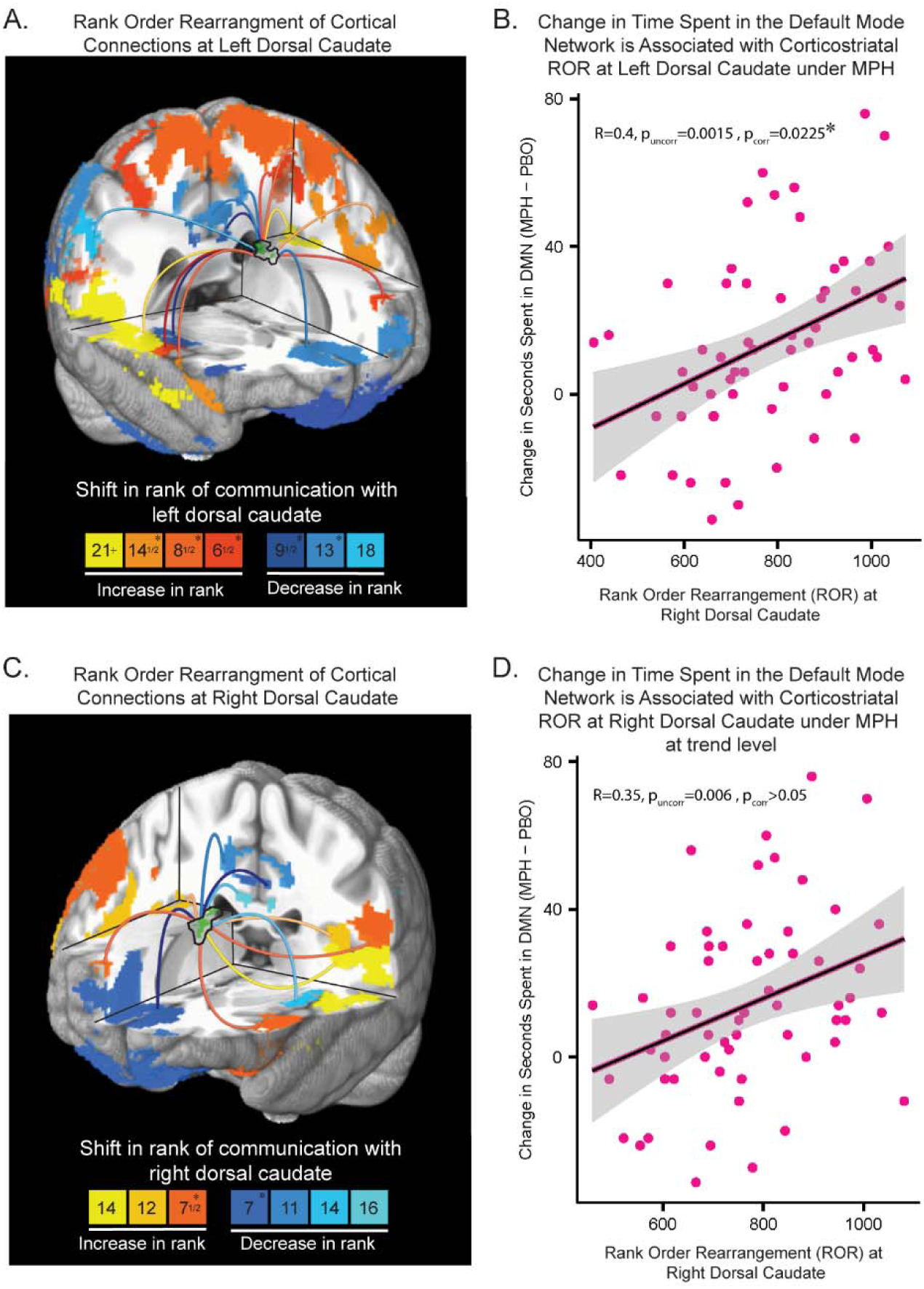
Methylphenidate-Induced Changes in the Relative Strength of Connectivity Between Cortical Regions and the Dorsal Caudate is Associated with More Time Spent in the Default Mode Network under Methylphenidate. A and C. ROR at left dorsal caudate (A) and right dorsal caudate (C), depicting absolute change in relative connectivity between multiple cortical regions and the dorsal caudate node under MPH compared to PBO: Green regions are the data-driven striatal nodes of significant MPH-induced ROR previously identified by a voxel-wise paired t-test. Cortical regions are color coded to depict how their relative connectivity strength with the specified caudate node, or ‘rank of communication’, shifts under MPH compared to under PBO. Regions shown shift at least 5 ranks. Color key at bottom indicates magnitude of change in rank where * indicates a mean magnitude of change for that color – this value only varies by ±2 values at maximum. Exact amount of change per significant cortical region is depicted in supplementary material figure S3. B and D. Correlations between ROR at left dorsal caudate (B) and at right dorsal caudate (D) and change in time spent (seconds) in the DMN under MPH compared to PBO. MPH: methylphenidate; PBO: placebo; DMN: default mode network; *p_corr_<0.05.

## Discussion

Methylphenidate increased the amount of time spent in the DMN and DAN states by increasing the frequency of transitions into both states. This finding is compelling given MPH’s known role in attention and the fact that both states play a role in different types of attention.

While the DMN plays a critical role in processes requiring internally focused attention such as episodic memory(37–40), the DAN is involved in the regulation of external attention, which is required to meet external task demands(37,41). Thus, even at rest, MPH facilitates dynamic transitions into attentional states, suggesting that MPH primes the brain to engage in both internal and external attentional demands. MPH also suppressed the amount of time spent in the FPN. Previous work has shown that the FPN facilitates goal-directed behavior via flexible switching between the DMN and the DAN to accomplish an internally or externally oriented task, respectively(37). Consistent with theories which posit MPH enhances cognitive efficiency(43), the suppression of time spent in the FPN coupled with enhanced entries into DMN and DAN may indicate that under MPH, less FPN-related effort is needed to facilitate goal-directed attention.

In a follow-up analysis, we confirmed that in addition to enhancing time spent in DMN and DAN, MPH strengthens static DMN-DAN anti-correlation. Stronger anti-correlation between the DMN and the DAN at rest is a known marker of healthy cognition(44), and it has been hypothesized that increased anti-correlation of these networks is driven by increased time spent in both brain networks, at opposing times(31). Here, we prove this conjecture that a stronger DMN-DAN anti-correlation is related to increased time in both DMN and DAN states. Our findings thus show that MPH drives changes in network temporal dynamics which explain known markers of cognitive enhancement identified by static functional connectivity measures.

Pharmacological challenge by HAL, in contrast, increased the amount of time in the SM-DMN brain state. Though it may appear counterintuitive that both MPH and HAL enhanced time in DMN-related states, the DMN and SM-DMN states are neurobiologically distinct representations of co-activated patterns(15). The enhancement of a SM-related network fits with known effects of antipsychotics on sensorimotor systems(42). Moreover, HAL is used as a first-line medication to treat schizophrenia, a disorder which exhibits altered temporal function of SM and DMN(14,45). The current finding in healthy controls that acute HAL enhances temporal function of two networks temporally disrupted in those with schizophrenia underscores the necessity of considering pharmacological impact of medication when analyzing temporal network function in clinical populations. This consideration may clarify mixed findings in the field(45).

Given that MPH and HAL have heightened locus of action at the striatum and act to modulate large-scale brain networks, we assessed how MPH and HAL impact corticostriatal function and whether this relates to drug-induced changes in network temporal dynamics. We obtained CSCPs, which aim to capture complex functional corticostriatal interactions by quantifying collective and relative connectivity between multiple cortical regions and striatal nodes, as AD, ROR, and ES. Challenge by HAL had no effect on CSCP measures. Challenge by MPH, in contrast, altered ROR but did not alter AD or ES. Previous work also has shown that each configuration property (AD, ROR, ES) is modulated independently and likely represents distinct neurobiological properties which may differentially shape striatal node function(24). We build on this to show that different pharmacological agents have distinct action on configuration profiles.

In altering ROR alone, MPH specifically acts to *reshuffle* the order of which corticostriatal connections are relatively greater or weaker at distinct striatal nodes. In doing so, MPH appears to rearrange the influence that different corticostriatal communications have(46) to ultimately contribute to altered brain function. It may be, at least in part, that MPH acting at the striatum significantly alters how cortical input is integrated in striatal cells(20) - though given MPH’s global action and effects on catecholamines broadly, future work is needed to disentangle this assertion. MPH-induced ROR occurs significantly at bilateral counterparts in the dorsal caudate and at the right and left nucleus accumbens and ventral caudate. Anatomically, these striatal nodes are engaged in bi-directional corticostriatal loops(47), and functionally, the dorsal caudate is tied to FPN while the ventral nodes identified here are linked to the DMN and limbic network(28).

Importantly, we provide novel evidence that MPH-induced corticostriatal ROR is tied to MPH-induced change in network temporal dynamics. The magnitude of ROR at the left and right dorsal caudate nodes are independently positively associated with network dynamic change; particularly related to enhancement of time spent in the DMN state. Translational research has established that the dopamine system and corticostriatal circuits, involving the dorsal caudate specifically, are implicated in addiction, ADHD, and the pharmacological action of MPH in the treatment of ADHD(48–51). More recent evidence has built on this historical understanding to illustrate that activity of neurocognitive networks, particularly DMN, is aberrant in these disorders(3,7,9,52,53). However, a gap remains in linking the often more pre-clinical body of circuit-based evidence to large-scale brain network function. Our findings reveal a link between MPH-induced changes in functional corticostriatal circuitry and temporal dynamic function of the DMN. These results may suggest that dopaminergic agonism alters how cortical connections with the dorsal caudate are prioritized, leading to changes in temporal dynamic function of brain networks, particularly the DMN. If this is true, it would expand on theory that *static* DMN functional connectivity is modulated by D2/D3 receptors(3) and suggest a specific locus and mechanism of regulation at the dorsal caudate, a striatal region functionally defined by its role in cognitive control(28,54).

### Limitations

A limitation of this present work is that baseline dopaminergic function was not assessed using PET. This may be relevant as others have shown that individual variance in dopaminergic function may impact the action of MPH(55,56). However, the current participants were healthy with no clinical appearance of dopaminergic disruptions. We also used a within-subjects design, which reduces the impact of individual variance. While we defined a relationship between temporal dynamics and corticostriatal configuration, we relied on correlational analyses and are unable to determine a causal relationship between these changes. As articulated in the discussion, we speculated that changes in corticostriatal interactions may drive the influence of MPH on temporal dynamic function; this hypothesis needs to be confirmed in future work. However, even in the absence of a causal link, we provide comprehensive evidence of MPH’s impact on temporal dynamics and related static resting state features that differ significantly from HAL.

### Conclusion

We find that pharmacological agents – MPH and HAL – have distinct impacts on network temporal dynamics and on corticostriatal configuration. Temporal dynamic findings suggest that MPH may prime the brain to engage in external or internal oriented attention, revealing a novel understanding of MPH action in alignment with the known impact of MPH on attention. Further, we show that while HAL does not alter corticostriatal configurations, MPH specifically alters ROR, acting to reshuffle the relative connectivity strength of multiple cortical regions with the striatum. The magnitude of this ‘reshuffling’, across the striatum and specifically at dorsal caudate is tied to MPH-induced change in brain network temporal dynamics, particularly the DMN. Thus, this work suggests that MPH-induced changes in dorsal caudate communication with the cortex may play a role in driving the time-varying engagement of the DMN and other large-scale brain networks, though future work is required to determine the causality of this association. Our findings ultimately uncover catecholamine-driven links between nuanced corticostriatal circuitry and large-scale brain network temporal dynamics, paving the way for a mechanistic understanding of the neurochemistry and neurocircuitry governing macro-scale brain network engagement.

## Data Availability

Data is not currently available for this data set but may become so upon reasonable request and regulatory approval.

## Acknowledgments

This work was supported by the National Institute on Drug Abuse Intramural Research Program We thank Dr. Blaise Frederick for his assistance in applying co-activation pattern analysis to this data.

