## Supplemental Information for "Catecholaminergic modulation of large-scale network dynamics is tied to the reconfiguration of corticostriatal connectivity"

**Supplementary Methods**

**Participants & Recruitment: supplementary methods**

This work was part of a larger trial evaluating genetic variance of the alpha 5 nicotinic subunit (SNP rs16969968) and included administration of nicotine patch. This was not an aim of the present study which did not include a focus on nicotine and instead focused on catecholaminergic manipulation specifically. It was verified that genetic variation in the SNP rs16969968 had no impact on the data.

**Exclusion Criteria**

Participants were required to be free of any lifetime history of substance use disorder and free of substance abuse for two years prior to participation as defined by DSM-IV diagnostic criteria. Exclusionary diagnoses also included but were not limited to psychiatric disorder, HIV positive status or neurological illnesses. Exclusionary medications included but were not limited to benzodiazepines, barbituates, anticonvulsants, antipsychotics, antidepressants, cold medicine and certain herbal medications (Kava, Gingko biloba). In addition, current use of strong or moderate CYP3A4 or 2D6 inhibitors was excluded as use may cause increased HAP concentration.

**Co-Activation Pattern Analysis**

**Supplementary Figure 1. 8 Brain states used in co-activation pattern analysis.**

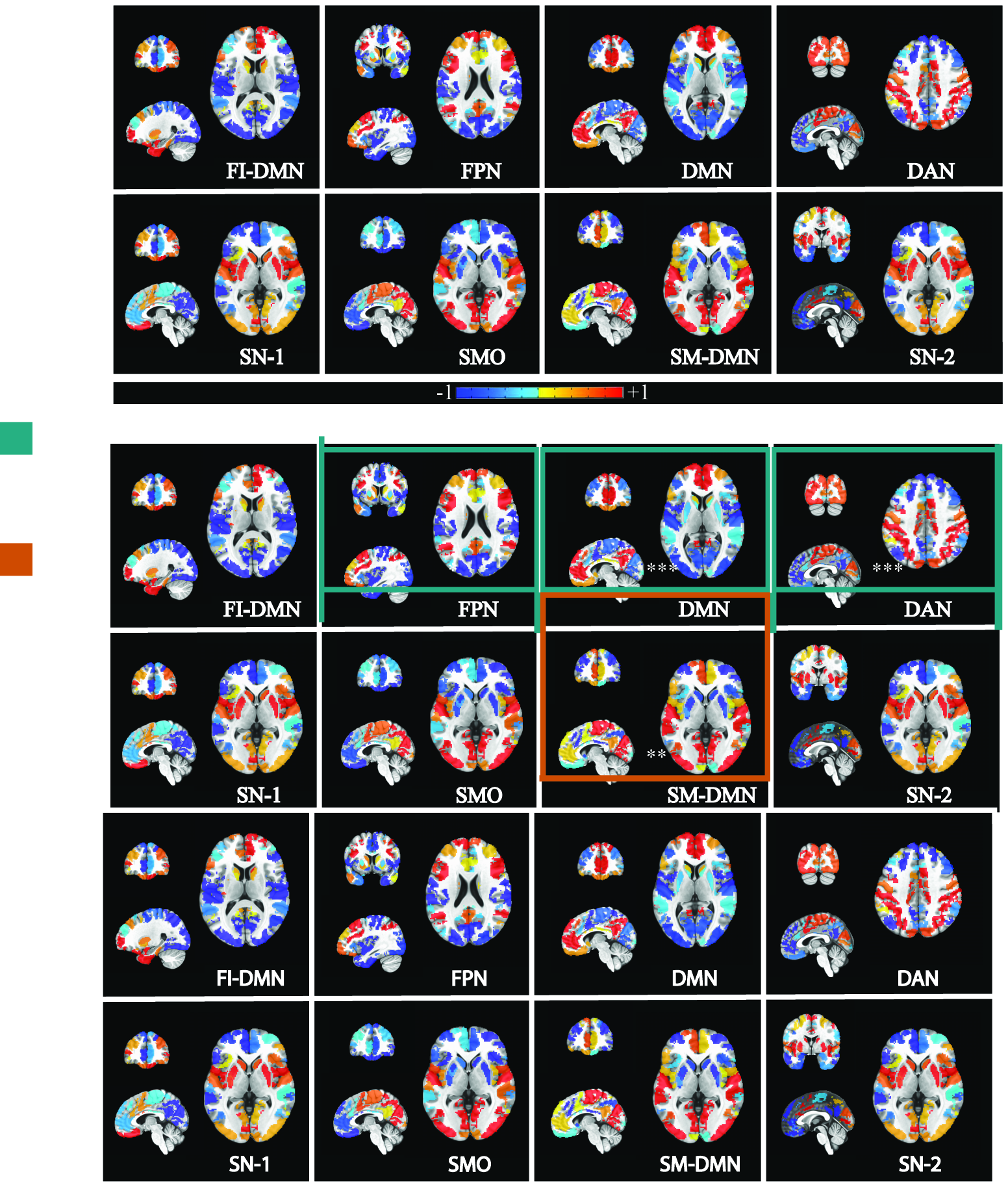

Supplementary Figure 1. 8 Brain states used in co-activation pattern analysis. -1 is most negative relative activation; +1 is most positive relative activation. FI-DMN: fronto-insular default mode network state; FPN: frontoparietal network state; DMN: default mode network state; DAN: dorsal attention network state; SN-1: salience network state 1; SMO: sensory motor occipital state; SM-DMN: sensory motor occipital default mode network state; SN-2 salience network state 2.

**Transitions Analysis**

To further bolster that the transitions were ‘meaningful transitions’ where the brain moved from sustained time in one state to sustained time in another state, we analyzed sustained transitions in R using timecourse files of each brain state assignment at each TR. We determined a sustained amount of time as more than 2 TRs in a row in the same brain state and calculated transitions between sustained states. To account for the possibility of noise between sustained transitions, we allowed a distance of 0 TRs or 1 TR between sustained states to detect a transition. For example, if the TR’s were repeatedly assigned to brain state 3, then for one TR to brain state 2, then repeatedly assigned to brain state 6, this was counted as a sustained transition from brain state 3 to brain state 6. As a follow up, we also calculated total transitions without taking into account these sustained transitions. These results largely replicated the results of the sustained transitions analysis.

**Corticostriatal Configuration Profiles Analysis**

To detect complex and nuanced changes in corticostriatal connectivity, we conducted connectivity profile analysis to compute corticostriatal configuration profile (CSCP) metrics. Time courses were extracted from each voxel of a striatal mask (Korponay et al., 2022) and from 53 cortical ROIs (used in CAPS states^1^; from Yeo et al., 2011; http://www.freesurfer.net/fswiki/CorticalParcellation_Yeo2011). The 53 ROIS are the bilateral and with-in 7-network combination of the cortical regions derived from the 17-network functional parcellation used to develop the co-activation patterns (8 brain states) applied here. 8 cortical regions did not have a contralateral counterpart and thus are unilateral in the analysis. We calculated the correlation between the time course of each cortical ROI and each striatal voxel, then performed Fisher’s R-to-Z transformation on correlation values.

These values Z-transformed correlation values were then back projected onto the striatum. This resulted in 53 striatal maps per subject and drug condition (53*3 per subject), containing the connectivity values at each voxel between that voxel of the striatum and each cortical ROI. These maps were then read into matlab 2022B to conduct the connectivity profile analysis.

Main analyses entailed the computation of three separate CSCP metrics, whose calculation is schematized in **Supplementary Figure 2** and detailed below, adapted from Korponay et al., (2022)^2^.

**Supplementary Figure 2. Computation schema of cortico-striatal profile configuration metrics.**

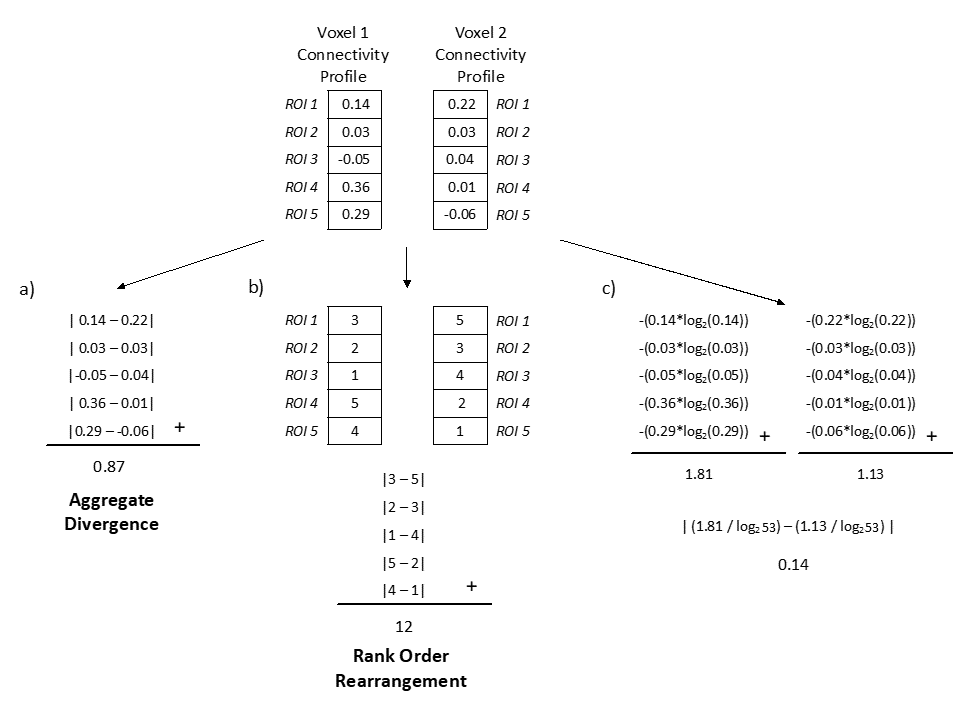

Supplementary Figure 2. Illustration of computation procedure for calculating the a) aggregate divergence, b) rank order rearrangement, and c) entropy shift between the connectivity profiles of two voxels with five cortical ROIs (our analysis used 53 ROIs). Numbers in the connectivity profile represent Z-scored functional connectivity between the voxel and each target ROI. Figure adapted from Korponay (2022).

*Aggregate Divergence*

To measure aggregate divergence (Figure S2a) between the connectivity profile under placebo compared to drug condition, we carried out the following procedure.

For each specific cortical region at a specific striatal voxel, we subtracted the z-score under placebo from z-score under drug condition. We then took the absolute value of this difference. Repeating this for all cortical regions at a specific striatal voxel, we next summed the absolute differences for all the 53 ROIs at the voxel. This produced the score of aggregate divergence at that voxel. We conducted this across all voxels for drug conditions compared to placebo. We also ran the same procedure comparing first and second half of placebo data. We then took the mean of aggregate divergence at all voxels per subject. This mean value was used in repeated measures ANOVA.

***Computing subject-wise AD value for methylphenidate condition***

**Value at each voxel**

$$\boldsymbol{AD voxel 1=}\sum_{\boldsymbol{i=ROI 1}}^{\boldsymbol{ROI 53}} \left| \boldsymbol{(}\boldsymbol{z score at voxel 1: Drug Condtion}_{\boldsymbol{i}}\boldsymbol{-}\boldsymbol{z score at voxel 1: Placebo Condition}_{\boldsymbol{i}}\boldsymbol{)} \right|$$

**Subject-wise score**

**SUBJECT AD = mean(AD voxel 1 _, … ,_ AD voxel n)**

*Rank Order Rearrangement*

Instead of constructing striatal voxel fingerprints using the Z-score connectivity values with each cortical ROI, here we used “rank order fingerprints” (Figure S2b) in which the value of each target ROI was its connectivity strength rank relative to the other ROIs in the fingerprint. As above, this procedure was carried out separately to compare fingerprint under methylphenidate and haloperidol to placebo, and to compare first half of placebo data to second half of placebo data.

After computing the rank order of each of the 53 cortical ROIs at each striatal voxel for each subject, we computed the absolute value of the difference between the rank order of each ROI at each voxel under drug compared to placebo and across the placebo session. The sum of these 53 absolute difference scores was then calculated to represent the magnitude of “rank order rearrangement” for each subject at each voxel. Similar to AD, we took the mean of rank order rearrangement at all voxels per subject. The mean value was used in repeated measures ANOVA.

***Computing subject-wise ROR value***

***Value at each voxel***

$$\boldsymbol{ROR voxel 1=}\sum_{\boldsymbol{i=ROI 1}}^{\boldsymbol{ROI 53}} \left| \boldsymbol{(}\boldsymbol{Rank at voxel 1: Drug Condtion}_{\boldsymbol{i}}\boldsymbol{-}\boldsymbol{Rank at voxel 1: Placebo Condition}_{\boldsymbol{i}}\boldsymbol{)} \right|$$

***Subject-wise score***

**SUBJECT ROR = mean(ROR voxel 1 _, … ,_ ROR voxel n)**

*Entropy Shift*

Using the Z-score connectivity voxel fingerprints, we calculated the entropy – a measure that indexes the uniformity of a distribution – of each voxel’s fingerprint for each subject:

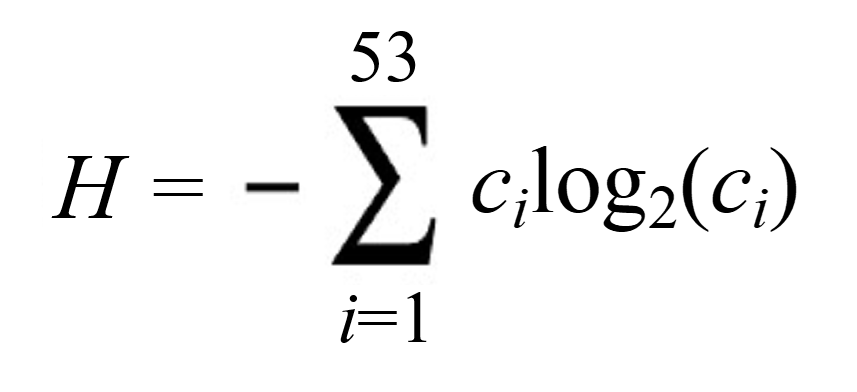

where *c_i_* is the magnitude of connectivity with the *i-*th cortical ROI. For each voxel we divided this measure by log_2_53 to normalize it and bind it to the interval [0,1]. Higher values of entropy represent connectivity profiles where connectivity strength is more evenly distributed across ROIs. Conversely, lower values of entropy represent connectivity profiles where connectivity strength is more concentrated with one or a few ROIs. The difference in each subject’s entropy at each voxel was than computed to represent the “entropy shift” between the drug conditions at each voxel. Like the other CSCP metrics, we took the mean of entropy shift at all voxels per subject. The mean value was used in repeated measures ANOVA.

*Localizing Striatal Nodes of Significant Difference in CSCP*

In configuration metrics (AD, ROR, ES) where there was a significant effect of drug determined by ANOVA and posthoc comparisons, we then aimed to identify specific areas of the striatum which were significantly altered by drug. To do so, we back projected subject-wise voxel-wise CSCP score (AD, ROR and/or ES) onto a map of the striatum for the drug condition of interest (drug-placebo). We also back projected subject-wise voxel-wise CSCP score (AD, ROR and/or ES) onto a map of the striatum for the PBO_firsthalf_-PBO_secondhalf_ condition. We then performed a voxel-wise paired t-test between significant drug-placebo scores and with-in subject placebo comparison scores using spm-12. Significant clusters were determined at p < .05 pFWE and at n > 10 voxels. Here, we determined that there was a significant effect of methylphenidate on ROR. Thus, we back projected 1) the voxel-wise score of MPH-PBO ROR onto a map of the striatum for each subject and 2) the voxel-wise score of PBO_firsthalf_-PBO_secondhalf_ for each subject and performed a voxel-wise paired t-test between these two striatal maps.

*Identifying Cortical Regions involved in Changing CSCPs*

To explore cortical regions and networks driving change in rank order rearrangement, we computed group-wise rank of cortical regions under placebo and MPH conditions. We detected significant change in rank per region as change greater than the mean change in rank under placebo. This method was adapted from the approach used by Korponay et al., (2022).

**Cortico-striatal configuration profile x temporal dynamics**

We tested whether, under the same drug, significant change in time spent in brain states is related to significant alterations in CSCP metrics. We calculated subject-wise scores of absolute change in time spent in states under drug. This measure was derived by summing the absolute value of change in time in each of the eight states (drug – placebo).

$$\boldsymbol{Absolute change in time spent in states=}\sum_{\boldsymbol{i=state 1}}^{\boldsymbol{state 8}} \left| \boldsymbol{(}\boldsymbol{time in state on DRUG}_{\boldsymbol{i}}\boldsymbol{-}\boldsymbol{time in state on PLACEBO}_{\boldsymbol{i}}\boldsymbol{)} \right|$$

We then tested the relationship between subject-wise score of significant CSCP metric and absolute change in time spent in states. Where there was a significant relationship, we aimed to confirm that this relationship, involving absolute change in time spent in *all eight* brain states, is specifically driven by change in time spent in brain states determined to be significantly modulated by drug. We confirmed this by comparing correlations where y varied as absolute change in time spent in: all eight states, states significantly altered by drug, states not significantly altered by drug, and x remained CSCP score.

$$\boldsymbol{Change in time spent in significant states=}\sum_{\boldsymbol{i=first significant state}}^{\boldsymbol{last significant state}} \left| \boldsymbol{(}\boldsymbol{time in state on DRUG}_{\boldsymbol{i}}\boldsymbol{-}\boldsymbol{time in state on PLACEBO}_{\boldsymbol{i}}\boldsymbol{)} \right|$$

$$\boldsymbol{Change in time spent in non-significant states=}\sum_{\boldsymbol{i=first non-significant state}}^{\boldsymbol{last non-significant state}} \left| \boldsymbol{(}\boldsymbol{time in state on DRUG}_{\boldsymbol{i}}\boldsymbol{-}\boldsymbol{time in state on PLACEBO}_{\boldsymbol{i}}\boldsymbol{)} \right|$$

For the co-correlation analysis, first, the following three correlations were measured:

3 Correlations:

1) Absolute change in all states x CSCP metric

2) Absolute change in time spent in significant states x CSCP metric

3) Absolute change in time spent in non-significant states x CSCP metric

Co-correlation analysis then tested that correlation 1 is significantly higher than correlation 3, and that correlation 1 does not significantly differ from correlation 2.

This confirmation allowed us parse out which networks are involved in the relationship between temporal dynamic network properties and functional corticostriatal circuitry.

*Localizing striatal nodes and identifying specific networks involved in relationship between CSCP score and change in dynamic network function*

We conducted further exploratory analysis to localize striatal nodes and individual networks involved in significant relationships between CSCP score and change in time spent in brain states. In contrast to previous analysis which used subject-wise scores of CSCP metric averaged across all striatal voxels, we computed subject-wise, node-specific scores of CSCP metrics for each of the significant striatal nodes, by averaging the CSCP scores across voxels of the specified cluster per subject. Nodes were defined by previous analysis, where we computed voxel-wise paired t-test on significant CSCP metrics to derive clusters (nodes) where drug-induced change in CSCP metric differs significantly from placebo. We then tested correlations between each node-specific CSCP score and the change in time spent in each significantly altered state. These correlations were corrected for multiple comparisons with the Bonferrroni correction.

This allowed us to localize striatal nodes and identify specific networks that contribute to the relationship between CSCP score and change in time spent in brain states.

**Cortico-striatal configuration profile x temporal dynamics methods with prior results**

Here, we determined the MPH alone significantly impacted DNF and CSCP metrics. Specifically, MPH specifically changed dynamics of FPN, DMN and DAN and significantly altered ROR. Thus, the following calculations were performed:

$$\boldsymbol{Absolute Change in time spent in brain states=}\sum_{\boldsymbol{i=state 1}}^{\boldsymbol{state 8}} \left| \boldsymbol{(}\boldsymbol{time in state on MPH}_{\boldsymbol{i}}\boldsymbol{-}\boldsymbol{time in state on PLACEBO}_{\boldsymbol{i}}\boldsymbol{)} \right|$$

***Change in time spent in FPN*, DMN*, DAN**** = |(time in FPN _MPH_) – (time in FPN_PLACEBO_)| + |(time in DMN_MPH_) - (time in DMN_PLACEBO_)| + |(time in DAN_MPH_) - (time in DAN_PLACEBO_)|

***Change in time spent in FI-DMN, SN-1, SMO, SM-DMN, SN-2*** = |(time in FI-DMN _MPH_) – (time in FI-DMN_PLACEBO_)|+ |(time in SN-1_MPH_) - (time in SN-1_PLACEBO_)| + |(time in SMO_MPH_) - (time in SMO_PLACEBO_)| + |(time in SM-DMN_MPH_) - (time in SM-DMN_PLACEBO_)| + |(time in SN-2_MPH_) - (time in SN-2_PLACEBO_)|

3 Correlations:

1) MPH-induced Total change in time spent in all states x subject-wise mean ROR

2) MPH-induced Change in time spent in FPN, DMN, DAN states x subject-wise mean ROR

3) MPH-induced Change in time spent in all states except FPN, DMN DAN states x subject-wise mean ROR

Co-correlation analysis tested that correlation 1 is significantly higher than correlation 3, and that correlation 1 does not significantly differ from correlation 2.

*Localizing relationship between MPH-Induced Changes in ROR and Change in DNF of FPN, DMN and DAN states*

We conducted 15 correlations comparing node-wise MPH-induced ROR scores (5 data drive nodes) and change in time spent in the FPN, DMN, and DAN (3 states; 5x3 = 15 correlation). Correlations were corrected for using Bonferroni correction (.05/15 = .003).

**Supplementary Results**

**Corticostriatal Configurations**

**Supplementary Table 1.** **Five clusters where MPH significantly induced higher magnitude of ROR compared to placebo.**

| Location | Central Coordinates | Cluster size (number of voxels) |
| --- | --- | --- |
| Right dorsal striatum | 20, -28, 20 | 40 |
| Left dorsal striatum | -20, -28, 22 | 50 |
| Right accumbens + right ventral caudate | 8, 6, -12 | 136 |
| Left accumbens | -6, 4, -10 | 59 |
| Left ventral caudate | -18, 24, -2 | 28 |

Location and size of five striatal clusters where MPH-induced ROR is significantly heightened compared to placebo (voxel size 2x2x2 mm).

**Cortical Regions Re-ordered In Rank of Connectivity with Striatum Under Methylphenidate**

Significant change in rank constituted a change in greater than or equal to 12 ranks as 11 ranks was the average shift in rank in placebo condition.

**Dorsal Striatum**

To enhance interpretability, here we group most findings from left and right dorsal caudate together – see figure S3 for how patterns vary laterally. All interpretation here refers to figure S3. In the salience network, all significantly altered regions showed lower rank under MPH. This included the insula, IPL, right anterior cingulate, dPFC, and the paramedian cortex (light green points). For the DMN (orange), generally more posterier regions of the DMN network (posterior cingulate, parahippocampal cortex, retrosplenial cortex), and also medial PFC, show a pattern of decrease in rank. However, ventral PFC of DMN exhibits an increase in rank (second ROI from right). Similarly for the FPN (lavendar), more posterior regions (IPL and posterior cingulate) decreased in rank while the lateral PFC, as well as the ventral precentral cortex of DAN (frontal regions), increased in rank. For lateral PFC, the shift in rank moves from mid-high to very high and is consistent bilaterally (left caudate: fifth ROI from left; right caudate: fourth ROI from left). A generalized conclusion from these findings may be that MPH enhances the influence of ventral and lateral frontal regions across DMN, DAN and FPN networks while diminishing the influence of more posterior regions of the networks and diminishing the influence of salience network regions. The particular enhancement of lateral PFC is interesting given the role of this region in task-switching, and our findings that MPH enhances time spent in opposing attentional states, DAN (external attention) and DMN (internal attention).

Somatomotor regions (dark green) and one visual region (yellow) also increased from very low ranks to low-mid level ranks.

**Ventral Striatum**

For the salience network (light green), almost all significantly altered regions showed lower rank under MPH, consistent with findings in the dorsal striatum. The regions which instead exhibit increased rank were the orbitofrontal cortex and ventral PFC at the left ventral caudate (fifth ROI from left; first ROI from right). The increase in rank of OFC was replicated in the OFC region of the limbic network, and for this OFC of the limbic network, the rank shifed from mid-high to very high (blue) and was consistent laterally for ventral caudate regions (left caudate: seventh ROI from left; right caudate: third ROI from left). Similar to dorsal caudate, ventral striatam also exhibited an increase in rank of lateral PFC of the FPN (lavendar; left caudate: 6^th^ ROI from left; right caudate: second ROI from left). For the nodes at right nucleus accumbens/ventral caudate and left nucleus accumbens, beyond the previously stated findings, network-based patterns cannot clearly be gleaned from the small number of significant ROIs which differ in rank. Broadly, MPH appears to potentially enhance the influence of OFC and lPFC at ventral striatum while diminishing the influence of multiple regions in the salience network.

For the striatal node at the left ventral caudate, there are a few more noteworthy patterns. The dorsal PFC of both the SN and FPN (second ROI from left) significantly decreased in rank. This is interesting in contrast to the finding that the lateral PFC of the FPN (purple, 6^th^ ROI from left) and ventral PFC of DMN and SN (first ROI from right) increased in rank. It illustrates a split in how MPH alters the influence of PFC at the left ventral striatum: ventral lateral PFC influence is strengthened while dorsal PFC influence is weakened. This split pattern goes further. A pattern emerged of decreased rank of regions in the DAN (dark purple) and of increased rank of regions in the DMN (orange). Taken together, it appears MPH may enhance the influence of reward, emotional and internal processing regions (DMN regions, OFC) and diminish the influence of more externally oriented attentional regions (DAN regions, dPFC) at the left ventral caudate.

**Supplementary Figure 3. Cortical Regions which are Significantly Re-ordered in Rank of Connectivity at Striatal Nodes by Methylphenidate**
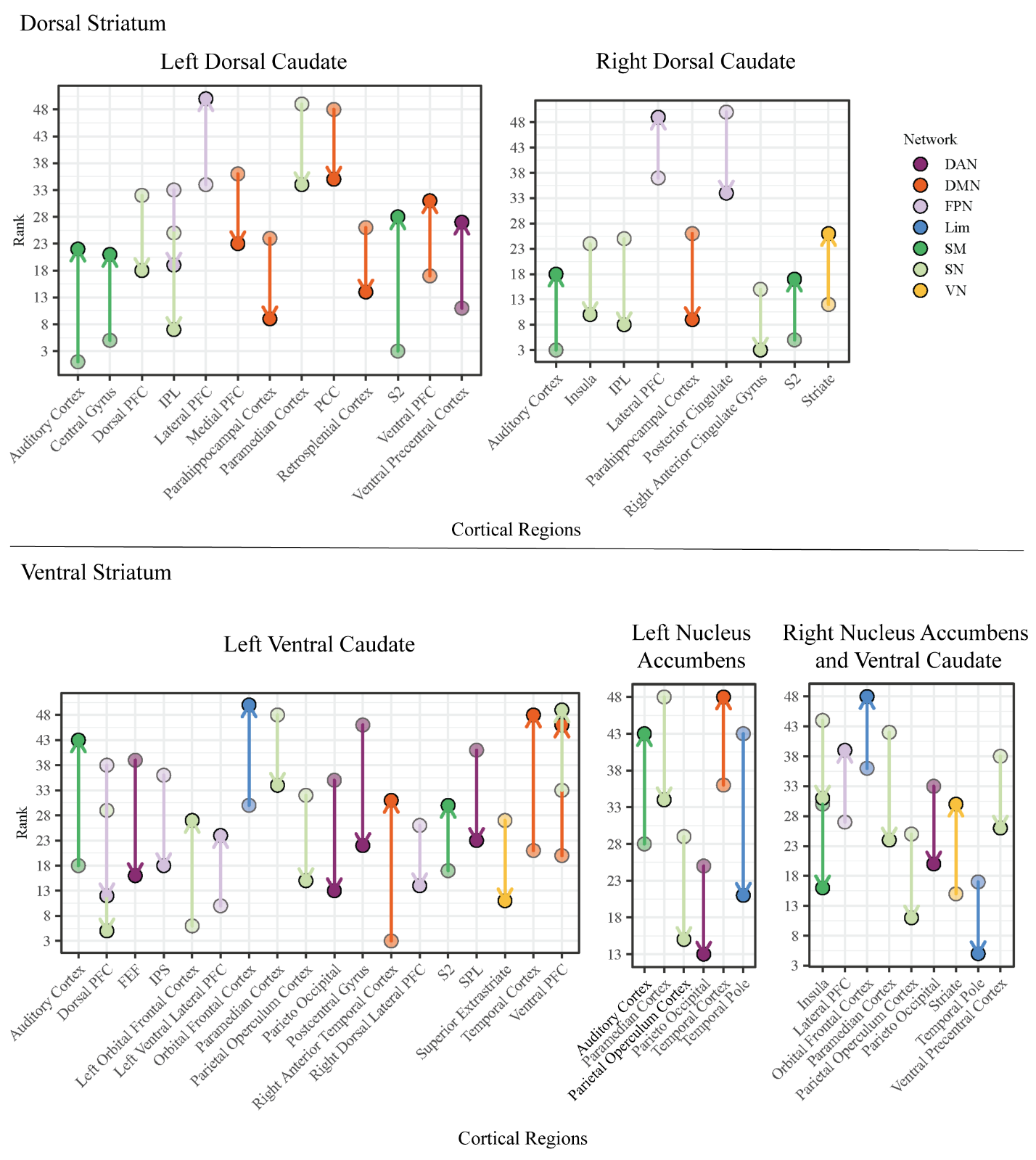

Supplementary Figure 3. Cortical rank order rearrangement under MPH compared to placebo separated by 7 networks at each of the five significant striatal nodes. 53 is highest rank, equivalent to strongest corticostriatal connection, 1 is lowest rank equivalent to weakest corticostriatal connection. Significant rearrangement is a change in greater than or equal to 12 ranks thus cortical regions shown here are at a minimum change in 12 ranks under MPH. MPH: methylphenidate. PFC: prefrontal cortex; IPL: inferior parietal lobe, PCC: posterior cingulate cortex, S2: secondary somatosensory cortex; FEF: frontal eye fields; IPS: intraparietal sulcus, SPL: superior parietal lobule.
